# Circulating microRNAs Predict Longitudinal Asthma Control and Treatment Response

**DOI:** 10.64898/2026.07.31.26359410

**Authors:** Parham Hadikhani, Alvin T Kho, Shraddha Piparia, Rinku Sharma, Scott T. Weiss, Michael McGeachie, Kelan G. Tantisira

## Abstract

**Background:** GINA-based clinical assessment of asthma control provides limited insight into the molecular mechanisms driving disease progression and treatment response. Circulating microRNAs (miRNAs) are implicated in immune regulation and airway remodeling, but their relationship to longitudinal, treatment-specific asthma control is not well characterized. We aimed to identify treatment-specific miRNAs associated with longitudinal asthma control and evaluate their ability to discriminate well-controlled from uncontrolled asthma.

**Methods:** Baseline serum miRNA sequencing data from 491 children in the Childhood Asthma Management Program (CAMP), a randomized trial of budesonide versus placebo, were analyzed, with GINA-based composite symptom scores assessed at baseline and 2, 4, 8, and 12 months. Cumulative link mixed models were fitted across 266 miRNAs to identify associations with longitudinal ordinal asthma control, adjusting for time, baseline status, and treatment. Random Forest classifiers were trained within each treatment group using Group K-Fold cross-validation. Pathway enrichment of validated miRNA targets was performed with DAVID.

**Results:** In the budesonide group, hsa-miR-1224-5p was associated with lower symptom severity and hsa-miR-199a-3p|hsa-miR-199b-3p with higher severity; both associations persisted at 12 months. The placebo group showed a broader pattern, with ten miRNAs associated with symptoms. Random Forest classifiers achieved mean AUC of 0.776 (budesonide) and 0.714 (placebo) for 12-month control status. Budesonide-associated targets were enriched for glucocorticoid-responsive and MAPK/Ras signaling, while placebo-associated targets showed broad enrichment for general regulatory processes.

**Conclusion:** Treatment-specific circulating miRNAs distinguish asthma control over time and implicate distinct signaling pathways, supporting their potential as complementary molecular markers for asthma monitoring in children.

## 1 Introduction

Asthma is a heterogeneous chronic respiratory disease characterized by variable airway inflammation, fluctuating symptoms, and diverse responses to therapy^11,2^. Despite the availability of effective controller medications, a substantial proportion of patients experience persistent symptoms or loss of disease control over time^3^. Clinical assessment of asthma control, commonly guided by Global Initiative for Asthma (GINA) criteria, relies on symptom frequency, rescue medication use, and functional limitations^4^. While these measures are clinically informative, they provide limited insight into the underlying molecular mechanisms driving disease progression and treatment response^2^. Identifying circulating biomarkers that reflect dynamic changes in asthma control could improve risk stratification, enable earlier intervention, and support more personalized management strategies^5^.

MicroRNAs (miRNAs) are small, non-coding RNA molecules that regulate gene expression post-transcriptionally and play critical roles in immune regulation, inflammation, and airway remodeling^6^. Circulating miRNAs are stable in peripheral blood and have emerged as promising non-invasive biomarkers for complex diseases, including asthma^7^. Prior studies have reported associations between specific miRNAs and asthma diagnosis, severity, or inflammatory phenotypes^8^; however, most investigations have been cross-sectional and focused on static disease states. As a result, relatively little is known about how baseline miRNA expression relates to longitudinal changes in asthma control or how miRNAs may differentially associate with clinical outcomes under distinct treatment conditions.

Longitudinal cohort studies are valuable because they track changes in the same individuals over time, both in molecular markers and clinical measures. For asthma, this approach can show whether baseline molecular profiles can predict future disease outcomes, instead of just reflecting current disease status^9^. Asthma control is naturally ordered, from well controlled to uncontrolled. To model disease progression accurately, we used statistical methods that preserve this ordinal structure and account for repeated measurements^10^.

Beyond association analyses, there is growing interest in leveraging molecular data to develop predictive models capable of classifying disease control status or anticipating treatment response. Machine learning approaches, particularly ensemble methods such as Random Forests, are well suited for high-dimensional miRNA data and can capture nonlinear relationships and interactions that may be missed by traditional regression models^11^. When combined with rigorous cross-validation and interpretable feature importance analyses, these models can complement inferential approaches by evaluating the practical utility of miRNA signatures as predictive tools^12^.

In this study, we leveraged longitudinal data from the Childhood Asthma Management Program (CAMP)^13^ to investigate the relationship between circulating miRNAs and asthma control over time. We pursued two complementary objectives. First, we used longitudinal ordinal mixed-effects modeling to identify miRNAs whose baseline expression is associated with subsequent asthma control severity and progression, while accounting for within-subject correlation and treatment effects. Second, we applied supervised classification analyses to assess the ability of significant identified miRNA panels to discriminate between well-controlled and uncontrolled asthma under different therapeutic contexts. By integrating longitudinal inference with predictive modeling, this work aims to identify miRNAs that are not only statistically associated with asthma outcomes but also clinically informative for forecasting disease control and treatment response.

## 2 Methods

### 2.1 Study Cohort

The Childhood Asthma Management Program (CAMP) was a four-year, multicenter, double-blind, randomized controlled trial (ClinicalTrials.gov Identifier: NCT00000575) of 1,041 children aged 5–12 years with mild-to-moderate persistent asthma, randomized to budesonide, nedocromil, or placebo^13^. We included 491 participants with baseline serum miRNA sequencing data; nedocromil-treated participants were excluded to allow direct budesonide-placebo comparison, and Partly Controlled participants were excluded from baseline analyses, yielding a baseline cohort of 190 (budesonide n=89; placebo n=101). Partly Controlled participants who transitioned to Uncontrolled or Well Controlled by follow-up were included at later visits (F12 n=232). Each site’s institutional review board approved the study; parents/guardians gave written informed consent and children gave assent. Full eligibility criteria and enrollment details are provided in Supplementary Methods.

### 2.2 Clinical Outcomes

Asthma control was assessed at baseline and 2, 4, 8, and 12 months (F0–F12) using a modified GINA composite score (range 0–3) summing three dichotomized symptom domains (SABA use, nocturnal awakening, exercise limitation), categorized as well controlled (0), partly controlled (1–2), or uncontrolled (3); a parallel continuous ordinal severity score (0–12) was also derived. Full scoring criteria and category definitions are provided in Supplementary Methods.

### 2.3 miRNA Sequencing and Processing

Small RNA sequencing was performed on baseline serum (Norgen Biotek Small RNA Library Prep Kit; Illumina NextSeq 500)^14^. Of 1,696 raw miRNAs, those with fewer than five counts in *≥*50% of samples were filtered, leaving 266 miRNAs, which were log_2_-transformed with a pseudocount of one.

### 2.4 Statistical and Machine Learning Analysis

A longitudinal long-format dataset linked baseline miRNA expression to time-specific GINA outcomes, treatment assignment, and time since baseline, using an available-case approach. Cumulative link mixed models (CLMMs; *ordinal* R package^15^) were fitted per miRNA, modeling GINA category as an ordinal outcome with time, baseline status, treatment, and a subject-specific random intercept as covariates; Wald p-values were corrected using the Benjamini-Hochberg false discovery rate procedure. Full model specification is provided in Supplementary Methods. Validated targets of significant miRNAs were retrieved via multiMiR (miRecords, TarBase, miRTarBase; *≥*2-database support)^16^ and evaluated for GO Biological Process, GO Molecular Function, and KEGG pathway enrichment using DAVID v6.8^17^ (FDR *≤* 0.10, minimum gene count *≥* 3).

Random Forest classifiers were trained to discriminate Well-Controlled from Uncontrolled asthma separately for budesonide (n=166, 437 observations) and placebo (n=243, 583 observations) cohorts, using CLMM-derived miRNA panels and baseline GINA score as a covariate. The pipeline used median imputation, feature standardization, and inverse-frequency class weighting. Performance was evaluated using Group *K*-Fold cross-validation (*K* = 5), grouped by participant identifier to prevent data leakage, reporting balanced accuracy, overall accuracy, weighted precision, recall, and F1-score, and area under the ROC curve. Permutation-based feature importance and chi-square analysis of aggregated confusion matrices were also performed. Full pipeline details and cross-validation rationale are provided in Supplementary Methods.

## 3 Results

### 3.1 Demographic and Clinical Characteristics of the CAMP Cohort

Demographic and clinical characteristics of the analytic cohort are presented in Table 1, stratified by treatment arm and asthma control status. After exclusion of nedocromil-treated and Partly Controlled participants, the baseline analytic cohort comprised 190 participants (budesonide: n=89, Uncontrolled n=68, Well Controlled n=21; placebo: n=101, Uncontrolled n=73, Well Controlled n=28).

**Table 1.** Demographic and clinical characteristics of the CAMP analytic cohort at baseline (F0) and 12-month follow-up (F12), stratified by treatment arm and asthma control status. Nedocromil-treated participants were excluded from all analyses. Partly Controlled participants were excluded from baseline analyses to enable a clinically meaningful binary comparison. Continuous variables are presented as mean ±SD. p-values compare Uncontrolled (UC) vs. Well Controlled (WC) within each treatment arm (Welch t-test for continuous; chi-square for categorical variables).

| Visit | Variable | Budesonide |  |  | Placebo |  |  |
| --- | --- | --- | --- | --- | --- | --- | --- |
|  |  | UC | WC | p | UC | WC | p |
| **F0** | *n* | 68 | 21 |  | 73 | 28 |  |
|  | Sex (Male) | 38 | 15 | 0.3104 | 39 | 21 | 0.0801 |
|  | Sex (Female) | 30 | 6 |  | 34 | 7 |  |
|  | Race (White) | 41 | 18 | 0.0688 | 65 | 25 | 1.000 |
|  | Race (Black) | 18 | 3 |  | 8 | 3 |  |
|  | Race (Hispanic) | 9 | 0 |  | 0 | 0 |  |
| | Age (years) | 9.25 ± 1.87 | 8.46 ± 2.35 | 0.1674 | 8.95 ± 2.06 | 8.23 ± 2.04 | 0.1179 |
| | BMI percentile | 64.74 ± 25.66 | 61.29 ± 28.18 | 0.6199 | 56.78 ± 28.67 | 59.05 ± 27.50 | 0.7238 |
| **F12** | *n* | 10 | 92 |  | 36 | 94 |  |
|  | Sex (Male) | 3 | 52 | 0.2062 | 19 | 60 | 0.3400 |
|  | Sex (Female) | 7 | 40 |  | 17 | 34 |  |
|  | Race (White) | 6 | 62 | 0.2954 | 29 | 78 | 0.9464 |
|  | Race (Black) | 4 | 20 |  | 7 | 16 |  |
|  | Race (Hispanic) | 0 | 10 |  | 0 | 0 |  |
| | Age (years) | 8.58 ± 2.33 | 9.16 ± 2.08 | 0.4718 | 8.90 ± 2.19 | 8.80 ± 2.18 | 0.8241 |
| | BMI percentile | 73.17 ± 24.28 | 62.71 ± 27.53 | 0.2267 | 56.36 ± 29.76 | 68.09 ± 28.06 | 0.0488 |

At baseline (F0), no statistically significant differences in sex, race, age, or BMI percentile were observed between Uncontrolled and Well Controlled participants within either treatment arm (all p > 0.05).

At the 12-month follow-up (F12), 232 participants had available outcome data classified as either Uncontrolled or Well Controlled (budesonide: n=102, Uncontrolled n=10, Well Controlled n=92; placebo: n=130, Uncontrolled n=36, Well Controlled n=94). The larger sample size at F12 compared with baseline reflects the inclusion of 135 participants who were classified as Partly Controlled at baseline and subsequently transitioned to either Uncontrolled or Well Controlled status by 12 months. The proportion of Well Controlled participants increased substantially in both treatment arms at F12 compared with baseline, consistent with overall improvement in asthma control over the study period.

No significant differences in sex, race, or age were observed between asthma control groups at F12 within either treatment arm (all p > 0.05). BMI percentile differed significantly between Uncontrolled and Well Controlled participants in the placebo group at F12 (56.36 ± 29.76 vs. 68.09 ± 28.06; p = 0.0488), but not in the budesonide group (73.17 ± 24.28 vs. 62.71 ± 27.53; p = 0.2267).

### 3.2 Treatment-Specific miRNA Associations with Symptom Severity

Fig. 1a presents the CLMM results for the budesonide group, revealing a relatively focused pattern of miRNA–symptom associations. Only a small number of miRNAs reached nominal statistical significance (*p <* 0.05). Among these, hsa-miR-199a-3p|hsa-miR-199b-3p displayed positive model coefficients, indicating associations with greater symptom severity, whereas hsa-miR-1224-5p showed a negative coefficient, suggesting an association with reduced symptom severity in the budesonide-treated context.

**Figure 1.**
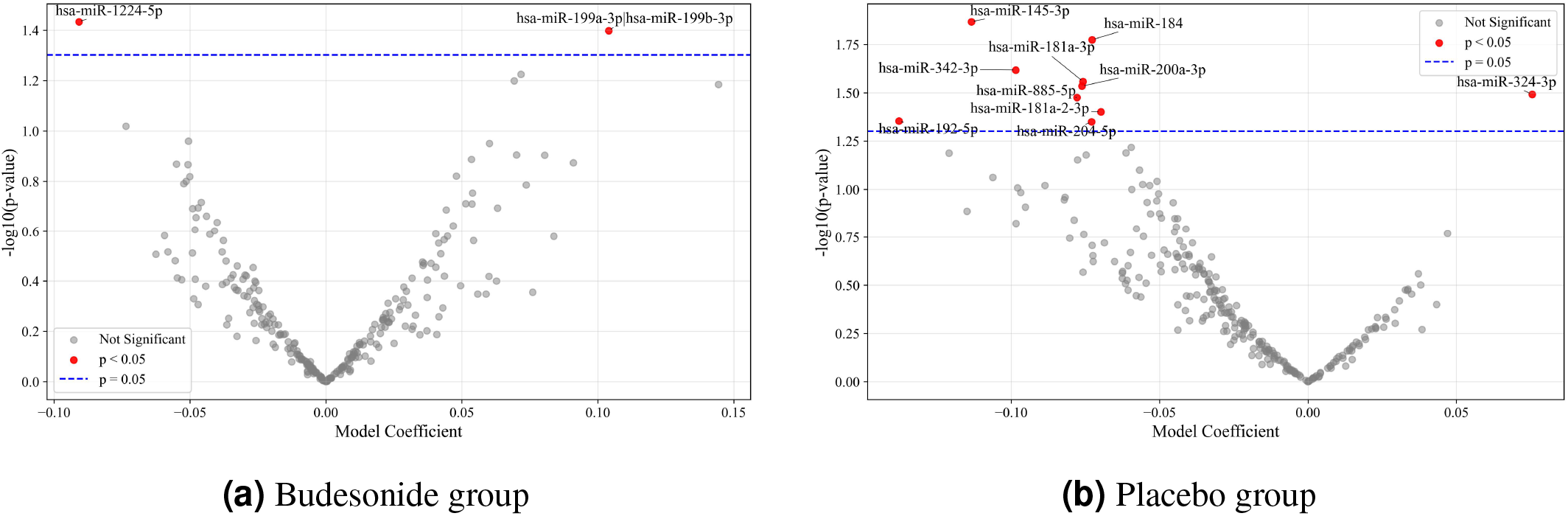
CLMM-derived miRNA–symptom associations stratified by treatment group. (a) Budesonide group and (b) placebo group. Each point represents an individual miRNA, with the x-axis indicating the regression coefficient and the y-axis representing *−*log_10_(*p*-value). The dashed horizontal line denotes the nominal significance threshold (*p <* 0.05). The placebo group shows a broader dispersion of nominally significant associations compared with the budesonide group.

In contrast, Fig. 1b shows that the placebo group exhibited a broader pattern of significant miRNA associations. Several miRNAs surpassed the nominal significance threshold. Most displayed negative model coefficients, indicating associations with reduced symptom severity over time, including hsa-miR-145-3p (the most statistically significant signal), hsa-miR-184, hsa-miR-342-3p, hsa-miR-181a-3p, hsa-miR-200a-3p, hsa-miR-885-5p, hsa-miR-181a-2-3p, hsa-miR-204-5p, and hsa-miR-192-5p. In contrast, hsa-miR-324-3p showed a positive model coefficient, indicating an association with increased symptom severity.

### 3.3 Baseline miRNA Expression Stratified by Concurrent Asthma Control Status

Fig. 2 shows mean baseline (F0) miRNA expression, averaged separately among participants classified as Well-Controlled versus Uncontrolled at each subsequent visit. Because control status can change between visits while miRNA expression was measured only at baseline, this analysis asks whether baseline expression consistently distinguishes participants by their current clinical status over the follow-up period, rather than depicting change in expression itself.

**Figure 2.**
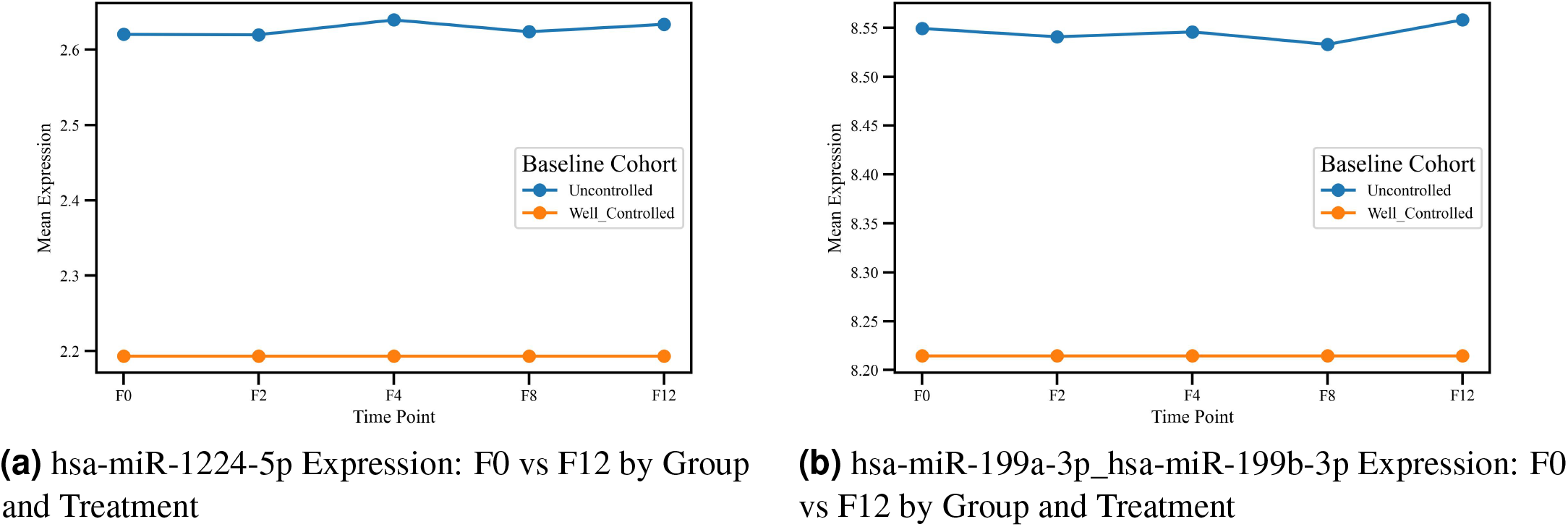
Mean baseline expression of budesonide-associated miRNAs, plotted at each follow-up visit according to participants’ concurrent asthma control status at that visit (not a repeated measurement — expression was assayed only at baseline). Group composition changes across visits as participants transition between control states. In the budesonide group, mean baseline hsa-miR-1224-5p expression was significantly higher among participants concurrently Well-Controlled (p = *p* = 2.73 × 10^*−*5^). In addition, hsa-miR-199a-3p|hsa-miR-199b-3p showed significant changes in Well-Controlled participants in both the placebo (*p* = 0.0014) and budesonide (*p* = 0.00148) groups. P values from two-sided Mann–Whitney U tests comparing baseline expression between concurrent control-status groups at each visit.

Distinct longitudinal patterns were observed across treatment and disease-control groups. In the budesonide group, participants who were Well-Controlled at F12 had significantly higher mean baseline hsa-miR-1224-5p expression than would be expected by chance (*p* = 2.73 × 10^*−*5^), an association not observed in the placebo group, suggesting the baseline signal is only informative in the context of corticosteroid treatment. Baseline hsa-miR-199a-3p|hsa-miR-199b-3p expression similarly distinguished Well-Controlled participants at F12 in both treatment groups.

In contrast, several miRNAs, including hsa-miR-184, hsa-miR-192-5p, hsa-miR-200a-3p, hsa-miR-204-5p, hsa-miR-145-3p, and hsa-miR-324-3p, showed the strongest baseline-expression separation when comparing participants who were Uncontrolled at F12 versus other outcomes, mainly in the placebo arm. This suggests baseline expression of these miRNAs is more strongly linked to persistent, untreated disease activity than to any change in the miRNA itself. This pattern is consistent with disease-related temporal changes in the absence of anti-inflammatory treatment. Other miRNAs, such as hsa-miR-885-5p, remained stable across groups, suggesting that the observed effects are specific rather than reflecting general time-related variation (Figs. S2-S6).

### 3.4 Longitudinal Asthma Control Trajectories

Longitudinal control trajectories (Fig. S7) indicated that among subjects Uncontrolled at baseline, budesonide-treated subjects showed rapid improvement by F2, whereas placebo-treated subjects followed slower improvement trajectories, with the number of Well-Controlled subjects exceeding the number of Uncontrolled subjects only at F12. Among subjects Well-Controlled at baseline, control was maintained in the budesonide group, whereas it was more variable in the placebo group.

### 3.5 Baseline miRNA Expression by Subsequent Asthma Control Transition

Fig. S8 compares baseline miRNA expression across subgroups defined by their control-status transition between F0 and F12 (e.g., remained Well-Controlled, or transitioned from Well-Controlled to Uncontrolled). Consistent with the CLMM analysis, baseline expression of these miRNAs differed according to the eventual transition pattern, indicating the baseline signal has prognostic association with future clinical trajectory. In particular, hsa-miR-1224-5p exhibited a clear increase among participants who remained Well-Controlled. In addition, hsa-miR-199a-3p|hsa-miR-199b-3p showed consistent changes across control transitions, with expression levels tending to differ between participants who remained Well-Controlled and those who transitioned to Uncontrolled states.

In contrast, several placebo-associated miRNAs showed expression changes primarily linked to disease-control transitions rather than treatment effects (Figs. S9–S13). These included hsa-miR-145-3p, hsa-miR-184, hsa-miR-342-3p, hsa-miR-181a-3p, hsa-miR-200a-3p, hsa-miR-885-5p, hsa-miR-181a-2-3p, hsa-miR-204-5p, hsa-miR-192-5p, and hsa-miR-324-3p. Across these miRNAs, expression patterns tended to vary between participants who remained in the same control category and those who transitioned between Well-Controlled and Uncontrolled states. However, the magnitude and direction of these changes were heterogeneous across miRNAs.

### 3.6 Distribution and Longitudinal Trends of Symptom Scores

Symptom score distribution (Fig. S14 was widely spread out and positively skewed at baseline (median 7), progressively decreasing throughout F12 (median 4), even as the proportion of patients with low symptom scores steadily increased.

### 3.7 Classification of Asthma Control Status Using miRNA Profiles

Table 2 shows the performance of Random Forest classifiers trained to distinguish Well-Controlled from Uncontrolled asthma using identified miRNA profiles. The values reflect mean performance aggregated across five held-out test folds, each from an independently trained Random Forest model. In the budesonide group, the average classifier achieved an overall accuracy of 0.723 and an AUC of 0.776, while in the placebo group, performance was similar, with an average accuracy of 0.693 and an average AUC of 0.714. Across both treatment groups, classification performance varied between groups, with the budesonide group having higher performance for Uncontrolled participants and the placebo group having higher performance for Well Controlled participants. In the budesonide group, the aggregate classifiers F1-score was 0.771 for Well-Controlled and 0.640 for Uncontrolled. In the placebo group, the corresponding aggregate classifiers F1-scores were 0.723 and 0.650. Weighted precision, recall, and F1-scores were similar between groups (approximately 0.69-0.78), indicating stable overall performance across all models generated through cross validation.

**Table 2.** Cross-validated classifier performance for GINA outcome.

| Treatment | Class | Per-Class |  |  | Overall |  |  |
| --- | --- | --- | --- | --- | --- | --- | --- |
|  |  | Precision | Recall | F1 | Accuracy | AUC | Weighted F1 |
| Budesonide | Well-Controlled | 0.806 | 0.736 | 0.767 | 0.724 | 0.79 | 0.725 |
|  | Uncontrolled | 0.630 | 0.695 | 0.656 |  |  |  |
| Placebo | Well-Controlled | 0.790 | 0.785 | 0.786 | 0.734 | 0.80 | 0.733 |
|  | Uncontrolled | 0.670 | 0.650 | 0.650 |  |  |  |

### 3.8 Confusion Matrix Analysis of miRNA-Based Classifier Performance

Fig. 3 shows the cross-validated confusion matrices for both treatment groups. In the budesonide group (Fig. 3b), 69.2% of Well-Controlled participants were correctly classified (211/305), while 79.5% of Uncontrolled participants were correctly identified (105/132). Misclassification occurred in both directions: 30.8% of Well-Controlled participants were predicted as Uncontrolled, and 20.5% of Uncontrolled participants were predicted as Well-Controlled. A similar pattern was observed in the placebo group (Fig. 3a). Well-Controlled participants were correctly classified in 71.2% of cases, while 66.8% of Uncontrolled participants were correctly identified. Misclassification of Uncontrolled participants (33.2%) was slightly higher than misclassification of Well Controlled participants (28.8%) in the placebo group.

**Figure 3.**
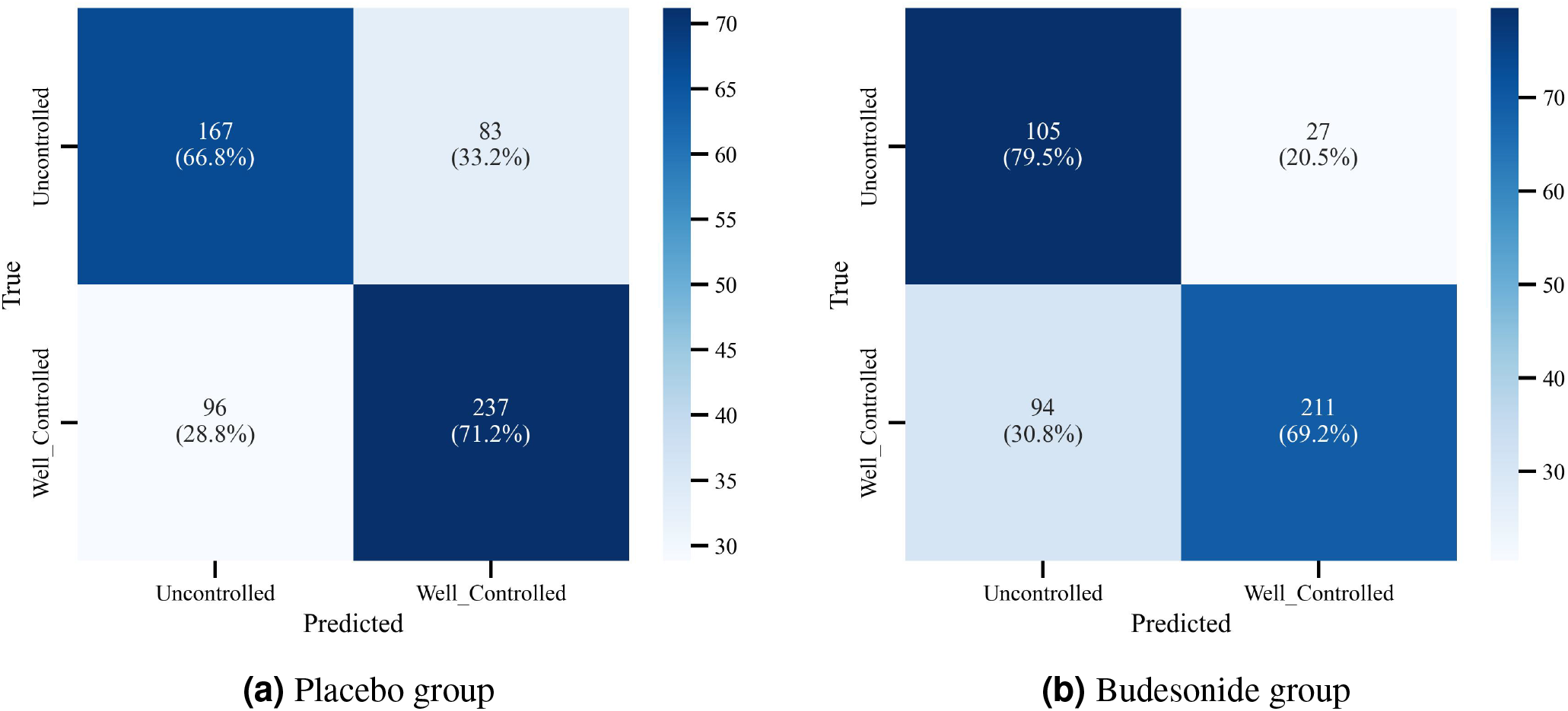
Cross-validated confusion matrices comparing observed GINA control status to predictions from the miRNA-based Random Forest classifier, in the placebo and budesonide treatment groups. Percentages indicate row-normalized classification accuracy within each true class.

Predicted labels were strongly associated with observed categories in both groups (*χ*^2^ *p* = 1.59 × 10^*−*20^ budesonide; *p* = 1.65 × 10^*−*19^ placebo), with standardized residuals confirming more correct classifications than expected by chance (Fig. S16).

### 3.9 Pathway Enrichment Analysis of Treatment-Specific miRNA Targets

Pathway enrichment of validated targets of budesonide-associated miRNAs revealed selective modulation of asthma-relevant signaling networks (Fig. 4a). The top-ranked pathway was Endocrine resistance, reflecting glucocorticoid receptor sensitivity (*−* log_10_(BH FDR) *≈* 5.15), followed by Nucleus with the largest gene set, highlighting transcriptional regulation (*−*log_10_(BH FDR) *≈* 4.6). Canonical inflammatory and remodeling pathways, including MAPK signaling, Ras signaling, EGFR tyrosine kinase inhibitor resistance, and TNF signaling, were also enriched (*−*log_10_(BH FDR) *≈* 4.0–4.5), consistent with budesonide’s known anti-inflammatory and airway remodeling effects. Ubl conjugation was moderately enriched, indicating involvement of post-translational modifications in pathway regulation.

**Figure 4.**
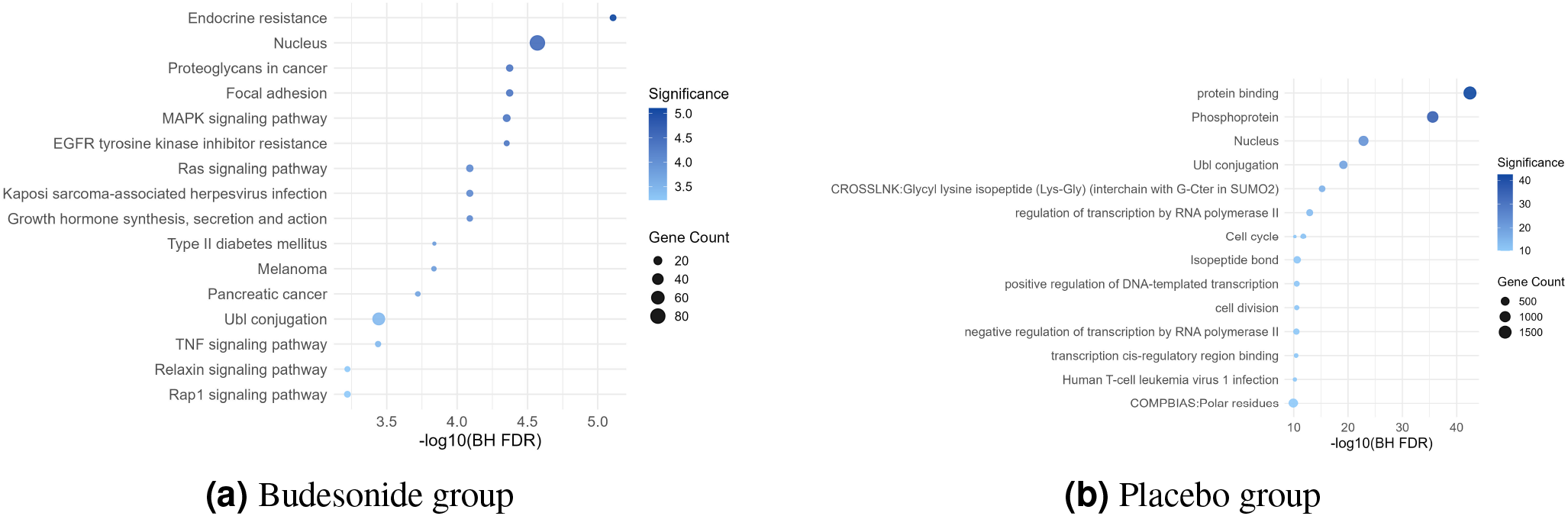
Pathway enrichment of validated targets of treatment-associated miRNAs. Targets were identified using multiMiR (miRecords, TarBase, miRTarBase) with at least two-source support. Enrichment was performed with DAVID v6.8 (GO Biological Process, GO Molecular Function, KEGG), using BH FDR correction (FDR *≤* 0.10, minimum 3 genes). Panels show (a) budesonide and (b) placebo. The x-axis shows *−* log_10_(BH FDR), and bubble size indicates the number of mapped genes.

In contrast, placebo-associated miRNA targets were broadly enriched for general cellular and regulatory processes rather than discrete signaling cascades (Fig. 4b). The most significant terms included protein binding and Phosphoprotein (*−* log_10_(BH FDR) *≈* 42 and 35, respectively), alongside Nucleus and Ubl conjugation (*−*log_10_(BH FDR) *≈* 20–22). These results suggest widespread activation of phosphorylation-dependent signaling, transcriptional regulation, and post-translational modifications in the absence of budesonide.

## Discussion

In this study, we analyzed treatment specific miRNA patterns associated with asthma control using longitudinal data from the CAMP cohort. By combining statistical modeling, longitudinal analysis, and machine learning, we identified groups of miRNAs linked to symptom severity, asthma control changes over time, and treatment exposure.

Baseline demographic analysis showed small differences between asthma control groups in sex and age. Participants with uncontrolled asthma at baseline included a higher proportion of males and were slightly older on average. However, these differences were not present at the 12 month follow up, indicating that demographic and clinical characteristics were generally similar between groups during the study period.

At the clinical level, the CAMP cohort showed an overall improvement in asthma control during the 12 month follow up period. The proportion of participants classified as well controlled increased over time, and symptom scores generally decreased. However, there remained considerable variability across individuals, indicating heterogeneous clinical trajectories.

The CLMM analysis identified different sets of miRNAs associated with symptom severity in the budesonide and placebo groups. In the budesonide group, two miRNA signals showed significant associations with symptoms. These were hsa-miR-1224-5p and the combined signal hsa-miR-199a-3p|hsa-miR-199b-3p. In contrast, a larger group of miRNAs was associated with symptoms in the placebo group. These included hsa-miR-145-3p, hsa-miR-184, hsa-miR-342-3p, hsa-miR-181a-3p, hsa-miR-200a-3p, hsa-miR-885-5p, hsa-miR-181a-2-3p, hsa-miR-204-5p, hsa-miR-192-5p, and hsa-miR-324-3p. Most of these miRNAs were associated with lower symptom severity over time, while hsa-miR-324-3p showed the opposite pattern and was associated with higher symptom severity. A larger number of miRNAs were associated with symptoms in the placebo group compared with the budesonide group. This pattern may indicate that anti-inflammatory treatment reduces molecular variability related to symptom severity.

Longitudinal expression analysis provided additional information about how these miRNAs changed over time. In the budesonide group, hsa-miR-1224-5p increased between baseline and follow up among participants who achieved or maintained well controlled asthma. The combined hsa-miR-199a-3p|hsa-miR-199b-3p signal also showed consistent changes across visits. These patterns suggest that these miRNAs may be related to treatment response or improved disease control during corticosteroid therapy.

Several miRNAs identified in the placebo group showed expression changes mainly among participants with uncontrolled asthma. These included hsa-miR-184, hsa-miR-192-5p, hsa-miR-200a-3p, hsa-miR-204-5p, hsa-miR-145-3p, and hsa-miR-324-3p. Other miRNAs such as hsa-miR-342-3p, hsa-miR-181a-3p, hsa-miR-181a-2-3p, and hsa-miR-885-5p showed more variable expression patterns across control transitions. These results suggest that placebo associated miRNAs may reflect broader changes related to disease activity rather than treatment effects.

Machine learning analysis further showed that miRNA profiles can help distinguish between well controlled and uncontrolled asthma. Random Forest models achieved moderate classification performance with area under the curve values of 0.776 for the budesonide group and 0.714 for the placebo group. Classification performance varied between groups, with the budesonide group showing higher sensitivity for Uncontrolled participants and the placebo group showing higher precision for Well Controlled participants. One possible explanation is that uncontrolled asthma may involve multiple biological mechanisms, which can lead to more heterogeneous molecular patterns. Confusion matrix and chi square analyses also showed a strong relationship between predicted and observed asthma control categories. These results suggest that circulating miRNAs contain information related to both asthma control and treatment exposure. Circulating miRNAs may reflect multiple biological processes involved in asthma, including airway inflammation, tissue remodeling, and treatment response. The budesonide associated miRNAs appear to reflect treatment related changes in disease regulation, while the placebo associated miRNAs capture broader patterns related to disease variability.

Budesonide treatment selectively modulates asthma-relevant signaling pathways, targeting key mediators of inflammation and airway remodeling. The MAPK pathway, including ERK1/2, JNK, and p38 MAPK, is central to propagating inflammatory signals in airway smooth muscle and epithelial cells, promoting cytokine production, leukocyte recruitment, and structural remodeling. Budesonide attenuates these effects, suppressing MAPK-driven inflammation and remodeling^18^. Ras GTPases act upstream of MAPK cascades, integrating extracellular inflammatory cues into intracellular responses; budesonide’s modulation of Ras-MAPK signaling may further limit eosinophilic inflammation and airway structural changes^19^. TNF signaling intersects with MAPK pathways, influencing glucocorticoid receptor function and corticosteroid responsiveness, highlighting budesonide’s role in restoring anti-inflammatory control in severe asthma^20^. Similarly, EGFR signaling, though not a classical inflammatory pathway, contributes to epithelial and smooth muscle remodeling by transactivating Ras/MAPK cascades. Its inhibition by budesonide may reduce airway hyperreactivity and mucus production^21^. Collectively, these findings underscore budesonide’s targeted suppression of convergent pro-inflammatory and remodeling networks.

In placebo-treated patients, there was broad activation of fundamental cellular processes, notably phosphorylation-mediated signaling, which is central to asthma pathogenesis. Chronic allergen exposure enhances phosphorylation of glucocorticoid receptors at serine-226 and activates p38 MAPK, promoting airway inflammation and Th2 cytokine production, including IL-4, IL-5, and IL-6, and contributing to airway hyperresponsiveness and steroid resistance^22^. Nuclear transcriptional regulation further amplifies these inflammatory responses. IL-33, a cytokine with a nuclear structural domain, resides in epithelial and endothelial cells and functions as an “alarmin” upon cellular stress or allergen exposure. Chromatin-associated IL-33 activates ILC2s and drives production of Th2 cytokines such as IL-4, IL-5, and IL-13, thereby initiating and sustaining allergic airway inflammation^23^. Ubiquitin-mediated protein modifications, including ubiquitination and SUMOylation, also play a critical role in controlling airway inflammation by regulating the stability, activity, and signaling of proteins involved in Th2 differentiation, dendritic and B cell function, and mast cell activation. Specific ubiquitin ligases and deubiquitinating enzymes, including USP38, Cbl-b, and MARCH1, modulate cytokine signaling, NF-*κ*B activation, and STAT phosphorylation, while SUMOylation further regulates airway epithelial responses and inflammatory gene expression, highlighting Ubl conjugation as a central post-translational mechanism in asthma^24^. Finally, transcriptional regulation via RNA polymerase II and key transcription factors integrates these extracellular signals into gene expression programs that drive allergic inflammation. Cytokines such as IL-4 and IL-13 activate the JAK–STAT pathway, leading to STAT6 phosphorylation, nuclear translocation, and recruitment of RNA polymerase II to orchestrate Th2 inflammation, IgE production, eosinophil recruitment, and airway remodeling, with additional contributions from NF-*κ*B and GATA3^25^. Together, these processes illustrate that while some pathways overlap with those modulated by budesonide (e.g., nuclear processes and ubiquitin conjugation), their widespread engagement in placebo-treated patients emphasizes that ICS therapy selectively narrows disease-relevant signaling rather than broadly suppressing cellular function.

Several limitations should be considered. The analyses were performed within a single clinical trial cohort, and the findings should be validated in independent datasets. In addition, although the classification models performed reasonably well, prediction accuracy was moderate. Including additional molecular or clinical features may improve performance. Further experimental studies will also be needed to clarify the biological roles of these miRNAs in asthma.

## Conclusion

This study demonstrates that circulating miRNA profiles reflect both clinical asthma control and treatment context in a longitudinal pediatric cohort. Treatment-specific miRNA associations, stable longitudinal expression patterns, and reproducible classification performance collectively support the potential role of circulating miRNAs as molecular indicators of asthma disease activity. Pathway analyses further implicate signaling networks involved in inflammation, airway remodeling, and corticosteroid responsiveness, providing a mechanistic framework linking miRNA regulation to asthma pathobiology.

## Data Availability

The data used in this study are not publicly available due to restrictions on data sharing and institutional policies. Access to the data is restricted to authorized members of the research team.

## Acknowledgment

This work was supported by the National Institutes of Health (NIH) grants R01 HL162570, R01 HL161362, R01 HL155742, R01 HL177625, and K99 HL183694.

## Key Message

Circulating microRNAs measured at diagnosis carry treatment-specific information about future asthma control in children. hsa-miR-1224-5p and hsa-miR-199a-3p|hsa-miR-199b-3p tracked with inhaled-corticosteroid response and could serve as accessible, blood-based markers to help identify treatment response and guide monitoring in pediatric asthma.

