## Supplementary Materials for "Circulating microRNAs Predict Longitudinal Asthma Control and Treatment Response"

### 1 Supplementary Methods

#### 1.1 Study cohort: Childhood Asthma Management Program (CAMP)

The Childhood Asthma Management Program (CAMP) was a four-year, multicenter, double-blind, randomized controlled clinical trial (ClinicalTrials.gov Identifier: NCT00000575) involving 1,041 ethnically diverse children aged 5–12 years with mild-to-moderate persistent asthma[1]. Between December 1993 and September 1995, participants were enrolled at eight clinical centers. Eligibility criteria included the presence of asthma symptoms, use of an inhaled bronchodilator at least twice weekly or daily asthma medication, airway hyperresponsiveness to methacholine (provocative concentration $\leq$ 12.5 mg/ml causing a 20% reduction in $FEV_1$), and absence of other clinically significant conditions. Participants were randomized to receive daily budesonide, nedocromil, or placebo.

For the present study, 491 participants with available baseline serum samples and miRNA sequencing data were included. Individuals randomized to nedocromil were excluded to allow direct comparison between budesonide and placebo groups. Furthermore, participants classified as Partly Controlled at baseline were excluded from baseline analyses to enable a clinically meaningful binary comparison between Well Controlled and Uncontrolled asthma. After these exclusions, the baseline analytic cohort comprised 190 participants (budesonide: n=89; placebo: n=101). At follow-up visits, participants originally classified as Partly Controlled at baseline who transitioned to either Uncontrolled or Well Controlled status were included in follow-up analyses, resulting in a larger available sample at later time points (F12: n=232).

Each child provided assent, and a parent or guardian gave written informed consent. Each participating study center obtained approval by the site's institutional review board (IRB)[2].

#### 1.2 Clinical Outcomes and Asthma Control Definitions

Asthma control was evaluated using a modified GINA assessment based on three symptom domains collected at baseline and at follow-up visits occurring 2, 4, 8, and 12 months after enrollment: (i) symptomatic use of short-acting $\beta_2$-agonist (SABA), (ii) nocturnal awakening due to asthma, and (iii) exercise limitation attributable to asthma. Standard GINA guidelines incorporate a fourth domain (daytime symptoms); however, this domain was unavailable during longitudinal follow-up and was therefore excluded.

Each symptom domain was recorded as a five-level ordinal variable reflecting symptom frequency, ranging from "Never," "At least once, but not monthly," "At least once per month, but not weekly," "At least once per week, but not daily" (or nightly for nocturnal awakening), to "Almost daily" (or almost nightly for nocturnal awakening).

For GINA-based analyses, symptom frequencies were dichotomized to approximate established asthma control thresholds. Responses of never, at least once but not monthly, and at least once per month but not weekly were classified as controlled. Responses of at least once per week but not daily and almost daily were classified as uncontrolled. Note that this dichotomization approximates, but does not exactly replicate, standard GINA criteria.

At each time point (baseline, 2 months, 4 months, 8 months, and 12 months, termed here F0, F2, F4, F8 and F12, with "F" for "follow-up"), a composite GINA symptom score was calculated by summing the three binary symptom indicators (SABA use, nocturnal awakening, and exercise-induced symptoms), yielding a score ranging from 0 to 3. Composite scores were categorized as well controlled (score = 0), partly controlled (score = 1–2), or uncontrolled (score = 3).

In parallel, an ordinal symptom severity score was computed using the original five-level scale for each domain. The summed ordinal severity across the three domains yielded a continuous measure of overall symptom burden (range 0–12) at each time point.

##### *1.2.1 Sample sequencing*

Small RNA sequencing (RNA-seq) in CAMP has been described previously, but in brief: sequencing was performed on serum samples from 491 baseline samples in the CAMP cohort. Serum samples were stored at −80°C at the Channing Division of Network Medicine. Library preparation for small RNA sequencing was conducted using the Norgen Biotek Small RNA Library Prep Kit (Norgen Biotek, Thorold, ON, Canada), and sequencing was performed on the Illumina NextSeq 500 platform using established protocols[3].

##### *1.2.2 miRNA Expression Processing and Filtering*

Raw count data included 1696 miRNAs in the CAMP cohort. To remove lowly expressed miRNAs, we filtered out miRNAs with fewer than five raw counts in at least 50% of samples. After filtering, 266 miRNAs remained in CAMP. Filtered counts were rounded and $\log_2$-transformed after adding a pseudocount of one to stabilize variance and accommodate zero counts.

#### 1.3 Longitudinal Dataset Construction

miRNA expression and clinical phenotype data were aligned using unique participant identifiers, retaining only individuals present in both datasets. A longitudinal dataset was constructed in long format, with each participant contributing one observation per available visit. Each observation included baseline miRNA expression values, baseline GINA control status, the time-specific GINA outcome category, the ordinal symptom severity score, treatment assignment (budesonide or placebo), and the time elapsed since baseline. Missing data were handled using an available-case approach, retaining individuals with baseline measurements and at least one follow-up observation without imputation.

#### 1.4 Longitudinal Ordinal Mixed-Effects Modeling

To identify miRNAs associated with asthma control severity over time (Objective 1), cumulative link mixed models (CLMMs) with a logit link were fitted using the clmm function from the ordinal R package[4].

GINA asthma control category was treated as an ordinal outcome, preserving its natural clinical ordering.

Let $Y_{ij}$ denote the GINA control category for subject $i$ at time $j$, where

$$Y_{ij} \in \{1, 2, \ldots, K\}.$$

For each miRNA, a separate model was fitted:

$$\operatorname{logit}\{\Pr(Y_{ij} \le k)\} = \theta_k - \Big(\beta_{\text{miRNA}} \cdot \text{miRNA}_i + \beta_{\text{time}} \cdot \text{time}_{ij} + \beta_{\text{baseline}} \cdot \text{BaselineGINA}_i + \beta_{\text{treat}} \cdot \text{Treatment}_i + b_i\Big), \quad k = 1, \ldots, K-1.$$

Here, $\theta_k$ are threshold parameters, and $b_i \sim \mathcal{N}(0, \sigma_b^2)$ represents a subject-specific random intercept accounting for repeated measurements. Time was modeled continuously as time since baseline.

The proportional odds assumption implies constant covariate effects across thresholds. Models were estimated using maximum likelihood with adaptive Gauss–Hermite quadrature. Models were restricted to miRNAs with sufficient variability and adequate outcome representation. Non-convergent or unstable models were excluded. Wald test p-values were extracted, and multiple testing correction was performed using the Benjamini-Hochberg false discovery rate (FDR) procedure.

### 1.5 Pathway Enrichment Analysis

For miRNAs identified as significant in longitudinal analyses, their validated targets were retrieved using the multiMiR R package[5], integrating miRecords (v4), TarBase (v8), and miRTarBase (v7.0). Targets supported by at least two independent databases or experimental methods were retained. Gene symbols were mapped to Entrez Gene IDs using org.Hs.eg.db. Functional enrichment analysis was conducted using DAVID (v6.8)[6] with the Homo sapiens genome as background. GO Biological Process, GO Molecular Function, and KEGG pathway categories were evaluated. Significance was determined using Fisher's exact test with FDR $\leq 0.10$, minimum gene count $\geq 3$).

### 1.6 Classification Analysis

To evaluate the predictive utility of identified miRNA panels, supervised classification models were developed to discriminate well-controlled from uncontrolled asthma.

#### *1.6.1 Outcome Definition*

A binary outcome was defined by comparing Well Controlled and Uncontrolled asthma, as described in Section 1.1. Classification analyses utilized all available longitudinal observations (F0–F12), excluding Partly Controlled observations at each visit. Baseline GINA score was included as a covariate. Separate classification analyses were conducted for treatment-specific (budesonide: n=166 participants, 437 observations; placebo: n=243 participants, 583 observations) and pooled (n=409 participants, 1020 observations) cohorts.

#### *1.6.2 Feature Selection and miRNA Panels*

Classification models used predefined miRNA panels derived from statistically significant longitudinal findings and biologically supported candidates. Panels were evaluated separately within treatment arms and in pooled analyses.

##### 1.6.3 Machine Learning Pipeline

A standardized pipeline was implemented, including median imputation for missing values, feature standardization, and Random Forest classification. Random Forest models were chosen for their robustness to nonlinear relationships and interaction effects. Class imbalance was addressed using inverse-frequency class weighting, and hyperparameters were kept fixed across analyses to ensure comparability.

##### 1.6.4 Cross-Validation and Performance Evaluation

Performance was assessed using Group $K$-Fold cross-validation ($K = 5$) across all longitudinal observations (F0-F12), with participant identifier used as the grouping variable to ensure that all observations from a given participant were assigned exclusively to either the training or test set, preventing participant-level data leakage from repeated measurements. Preprocessing steps were performed within each training fold to prevent data leakage.

Given the relatively small sample sizes within treatment-specific subgroups, a single held-out test set would provide insufficient statistical power for reliable performance estimation. Group $K$-Fold cross-validation was therefore employed to maximize data utilization. Reported metrics represent mean performance across held-out folds and should be interpreted as estimates of expected generalization performance for this class of models, rather than the performance of a single final model.

Primary performance metric was balanced accuracy. Additional metrics included overall accuracy, weighted precision, weighted recall, weighted F1-score, and area under the ROC curve (AUC). Mean ROC curves were obtained by interpolating true positive rates across folds.

##### 1.6.5 Model Interpretation and Error Analysis

Permutation-based feature importance was computed within each fold and averaged across folds. Aggregated confusion matrices were analyzed using chi-square tests of independence. Standardized residuals were calculated to identify prediction patterns deviating from chance.

### Supplementary Figures

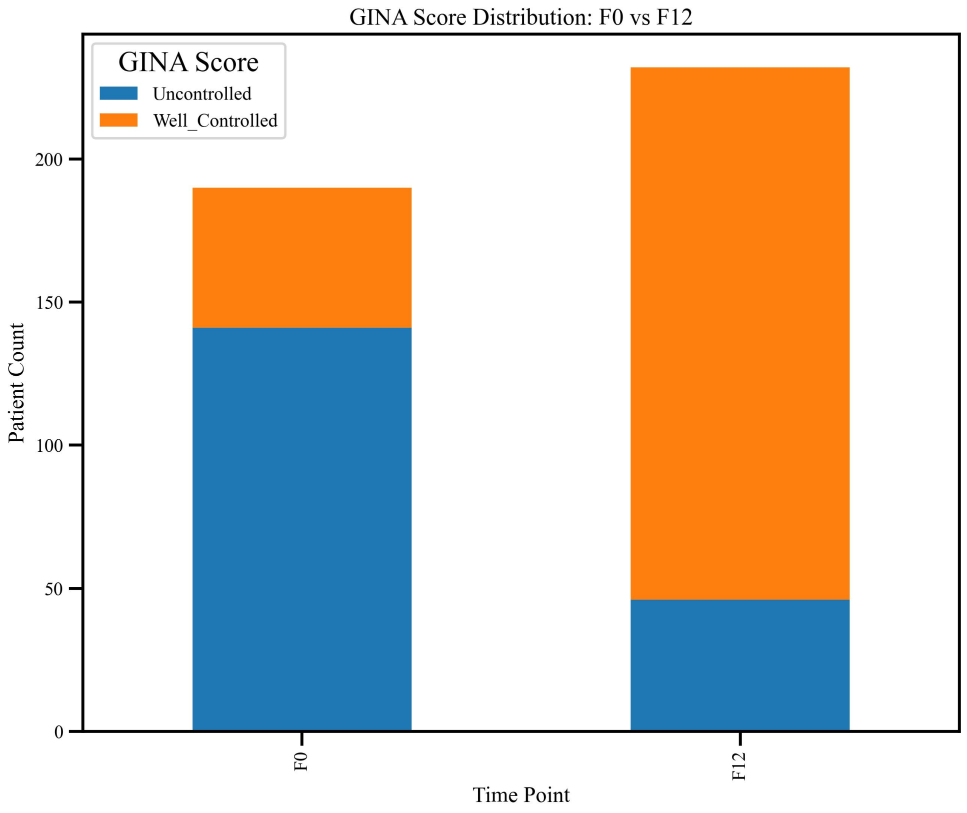


**SFigure 1.** Distribution of GINA asthma control categories at baseline and 12-month follow-up. Comparison of GINA outcome distributions at baseline (F0) and 12-month follow-up (F12) demonstrates a marked shift toward improved asthma control over time. At baseline, the cohort was predominantly classified as Uncontrolled, whereas at F12 the majority of participants were classified as Well-Controlled.

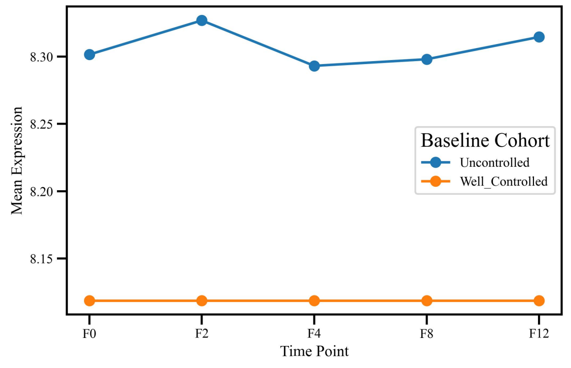


**(a)** hsa-miR-145-3p

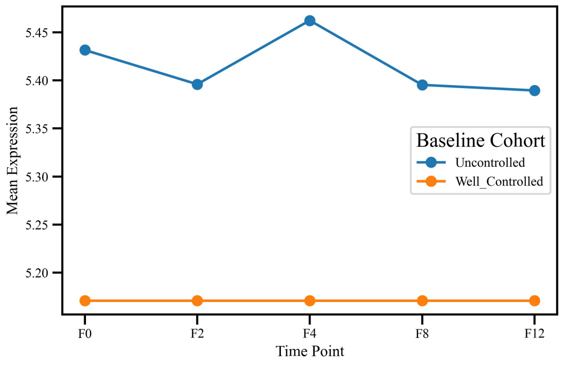


**(b)** hsa-miR-181a-2-3p

**SFigure 2.** Baseline expression of hsa-miR-145-3p and hsa-miR-181a-2-3p between baseline (F0) and follow-up (F12), stratified by asthma control status and treatment group (placebo or budesonide). Statistical comparisons were performed using two-sided Mann–Whitney U tests.

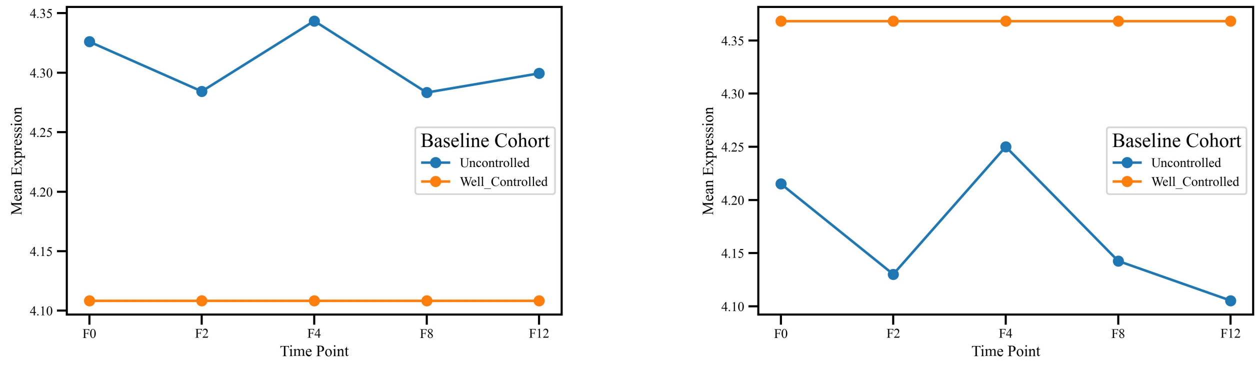


**(a)** hsa-miR-181a-3p

**(b)** hsa-miR-184

**SFigure 3.** Baseline expression of hsa-miR-181a-3p and hsa-miR-184 between baseline (F0) and follow-up (F12), stratified by asthma control status and treatment group (placebo or budesonide).

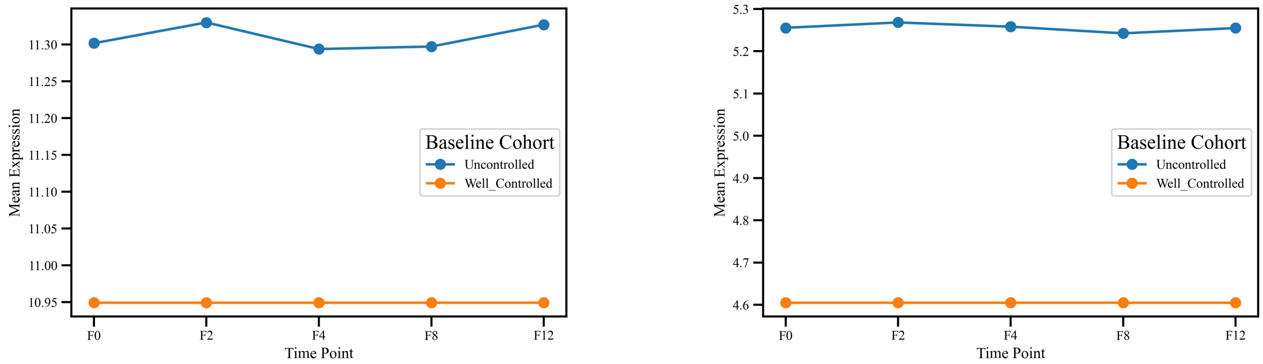


**(a)** hsa-miR-192-5p

**(b)** hsa-miR-200a-3p

**SFigure 4.** Baseline expression of hsa-miR-192-5p and hsa-miR-200a-3p between baseline (F0) and follow-up (F12), stratified by asthma control status and treatment group.

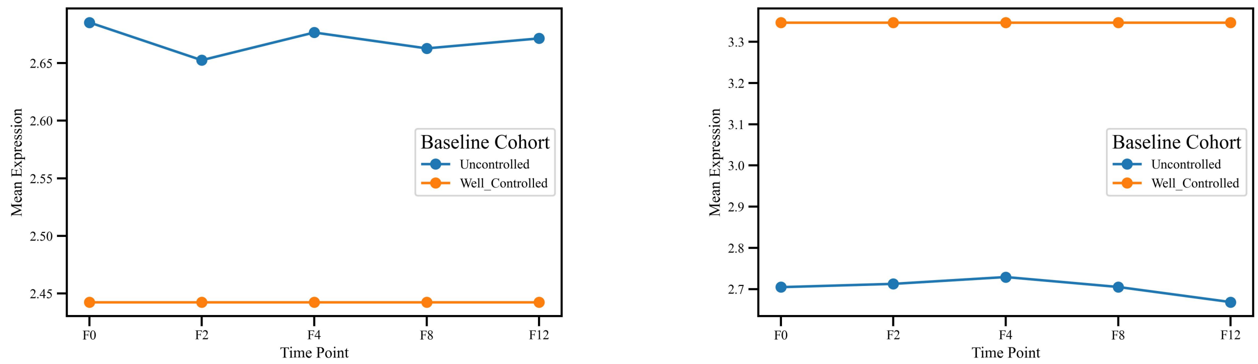


**(a)** hsa-miR-204-5p

**(b)** hsa-miR-324-3p

**SFigure 5.** Baseline expression of hsa-miR-204-5p and hsa-miR-324-3p between baseline (F0) and follow-up (F12), stratified by asthma control status and treatment group.

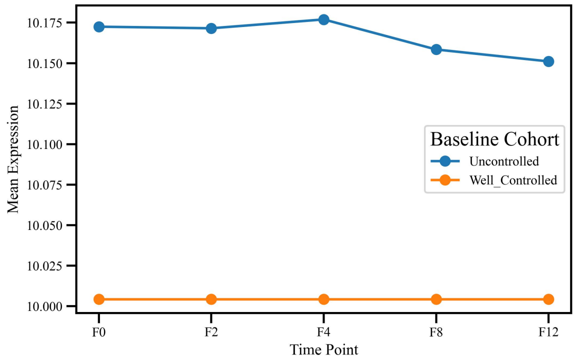


**(a)** hsa-miR-342-3p

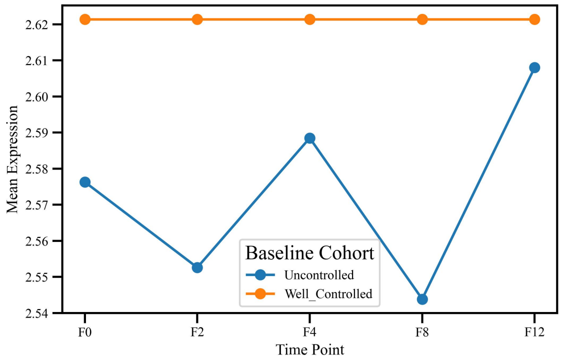


**(b)** hsa-miR-885-5p

**SFigure 6.** Baseline expression of hsa-miR-342-3p and hsa-miR-885-5p between baseline (F0) and follow-up (F12), stratified by asthma control status and treatment group.

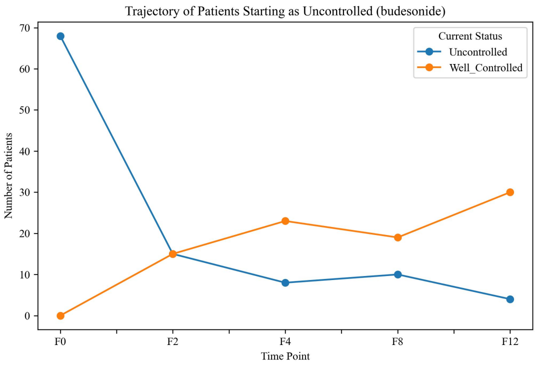


**(a)** Budesonide: baseline Uncontrolled

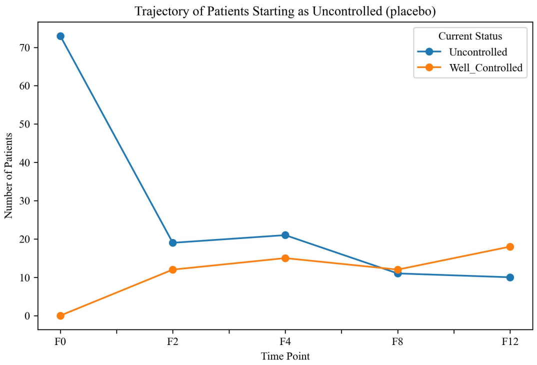


**(b)** Placebo: baseline Uncontrolled

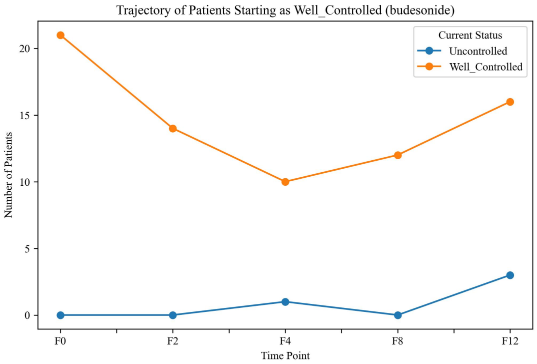


**(c)** Budesonide: baseline Well-Controlled

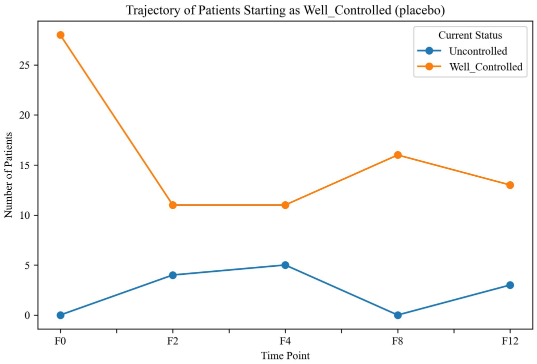


**(d)** Placebo: baseline Well-Controlled

**SFigure 7.** Longitudinal trajectories of asthma control across follow-up visits (F0, F2, F4, F8, F12) among participants classified as Uncontrolled or Well-Controlled at baseline, stratified by treatment group. Lines represent the number of participants observed in each asthma control category at each visit.

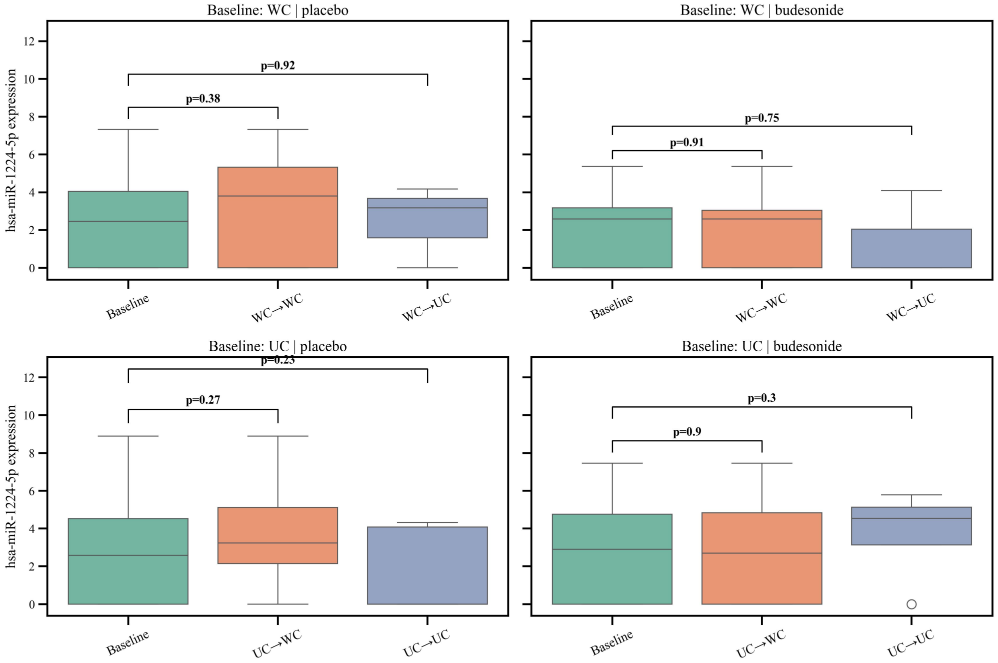


**(a)** hsa-miR-1224-5p Expression Transitions (F0 → F12) by Treatment

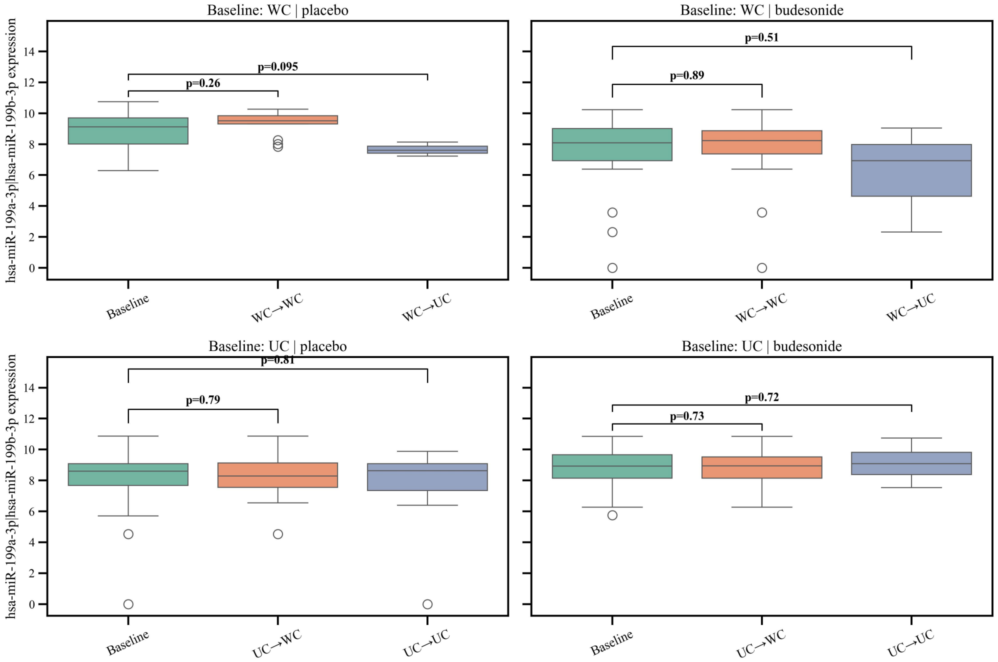


**(b)** hsa-miR-199a-3p / hsa-miR-199b-3p Expression Transitions (F0 → F12) by Treatment

**SFigure 8.** Baseline expression of budesonide-associated miRNAs across asthma control transition groups. Participants were grouped by baseline asthma control (well-controlled [WC] or uncontrolled [UC]) and their control-status transition from F0 to F12 (WC→WC, WC→UC, UC→WC, UC→UC). P values were calculated using two-sided Mann–Whitney U tests.

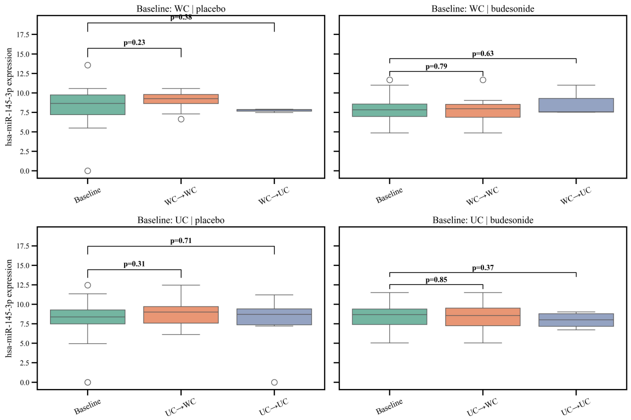


**(a)** hsa-miR-145-3p

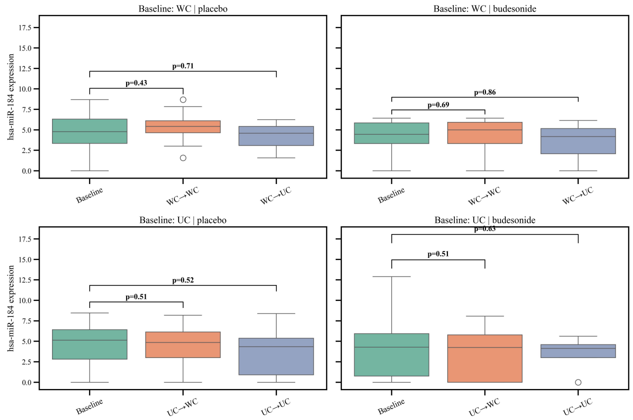


**(b)** hsa-miR-184

**SFigure 9.** Baseline expression of hsa-miR-145-3p and hsa-miR-184 across asthma control transitions. Expression levels are shown at baseline (F0) and follow-up (F12) among placebo-treated participants, stratified by baseline asthma control status (WC or UC) and subsequent control transitions (WC→WC, WC→UC, UC→WC, UC→UC). P values were calculated using two-sided Mann–Whitney U tests.

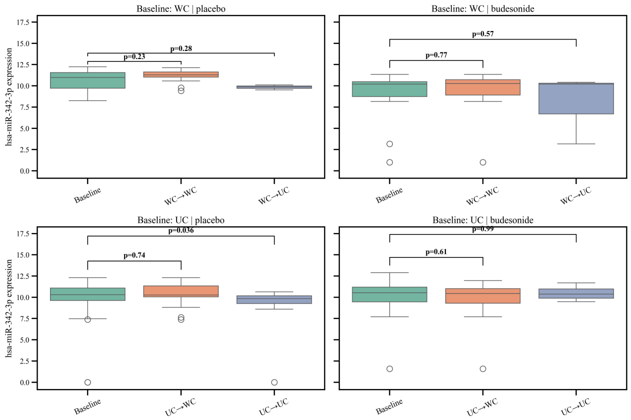


**(a)** hsa-miR-342-3p

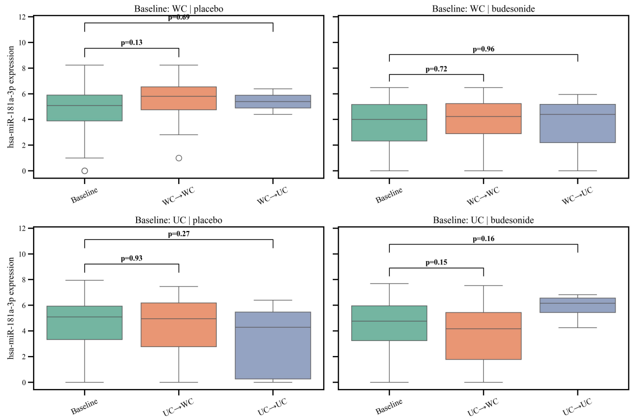


**(b)** hsa-miR-181a-3p

**SFigure 10.** Baseline expression of hsa-miR-342-3p and hsa-miR-181a-3p across asthma control transitions. Expression levels are shown at baseline (F0) and follow-up (F12) among placebo-treated participants, stratified by baseline asthma control status (WC or UC) and subsequent control transitions (WC→WC, WC→UC, UC→WC, UC→UC). P values were calculated using two-sided Mann–Whitney U tests.

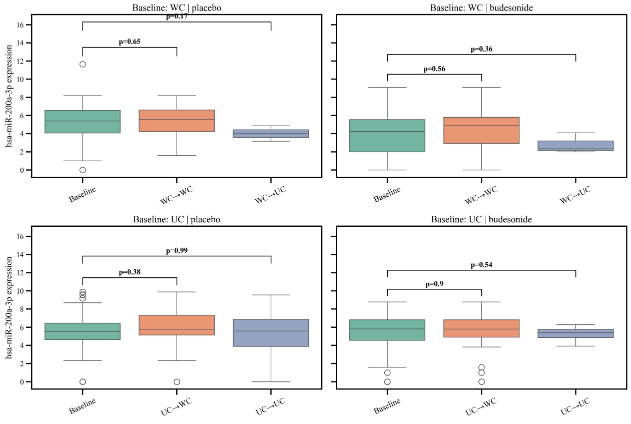


**(a)** hsa-miR-200a-3p

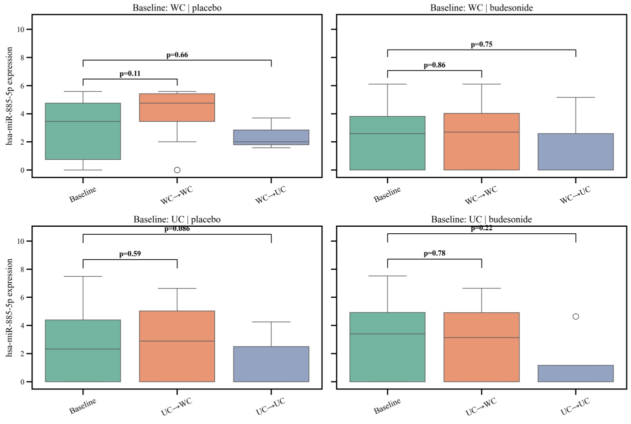


**(b)** hsa-miR-885-5p

**SFigure 11.** Baseline expression of hsa-miR-200a-3p and hsa-miR-885-5p across asthma control transitions. Expression levels are shown at baseline (F0) and follow-up (F12) among placebo-treated participants, stratified by baseline asthma control status (WC or UC) and subsequent control transitions (WC→WC, WC→UC, UC→WC, UC→UC). P values were calculated using two-sided Mann–Whitney U tests.

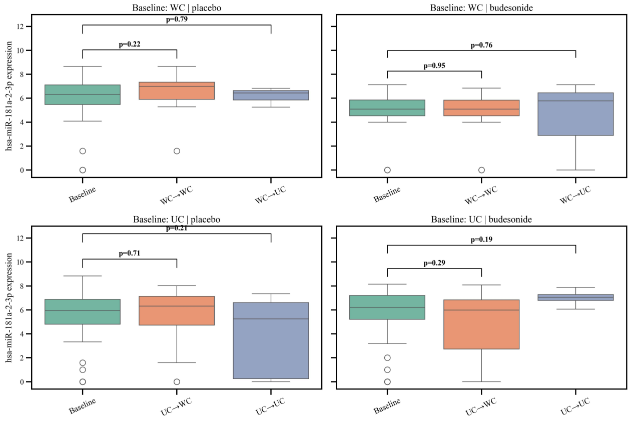


**(a)** hsa-miR-181a-2-3p

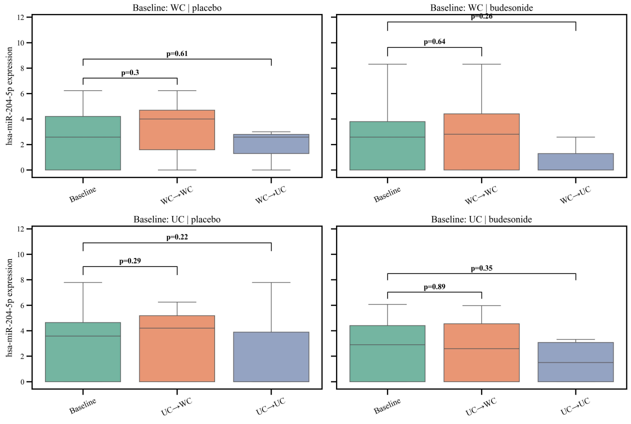


**(b)** hsa-miR-204-5p

**SFigure 12.** Baseline expression of hsa-miR-181a-2-3p and hsa-miR-204-5p across asthma control transitions. Expression levels are shown at baseline (F0) and follow-up (F12) among placebo-treated participants, stratified by baseline asthma control status (WC or UC) and subsequent control transitions (WC→WC, WC→UC, UC→WC, UC→UC). P values were calculated using two-sided Mann–Whitney U tests.

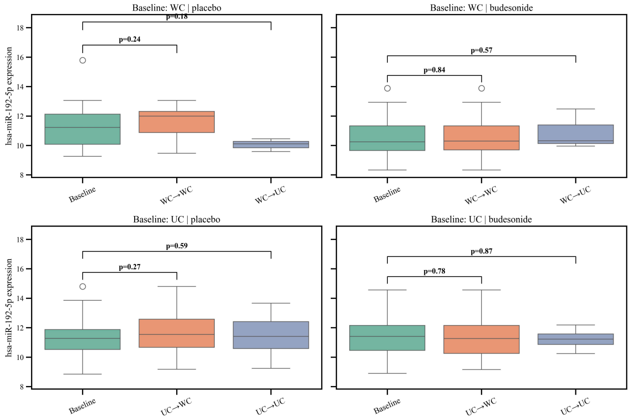


**(a)** hsa-miR-192-5p

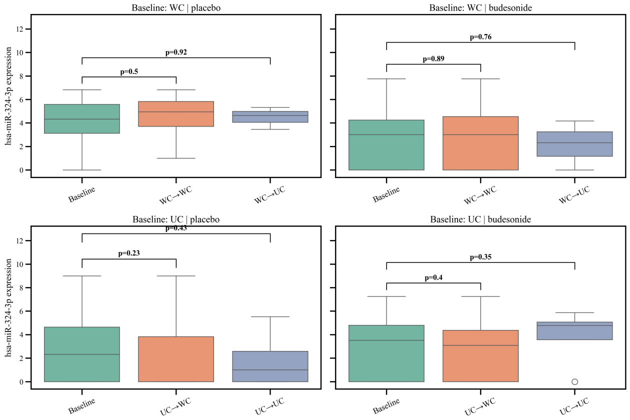


**(b)** hsa-miR-324-3p

**SFigure 13.** Baseline expression of hsa-miR-192-5p and hsa-miR-324-3p across asthma control transitions. Expression levels are shown at baseline (F0) and follow-up (F12) among placebo-treated participants, stratified by baseline asthma control status (WC or UC) and subsequent control transitions (WC→WC, WC→UC, UC→WC, UC→UC). P values were calculated using two-sided Mann–Whitney U tests.

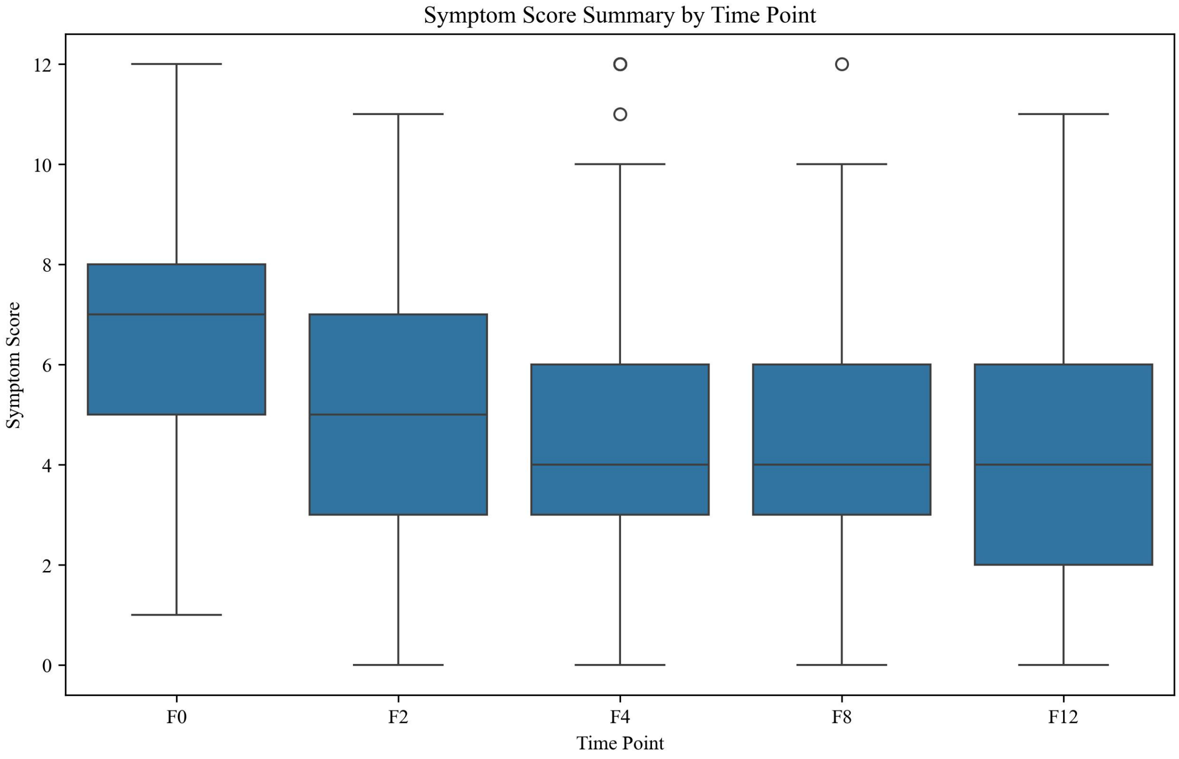


**SFigure 14.** Distribution of symptom scores across study visits. Boxplots show symptom score distributions at each visit (F0, F2, F4, F8, and F12). The center line represents the median, the box shows the interquartile range, and whiskers indicate the range excluding outliers. Points represent individual outliers.

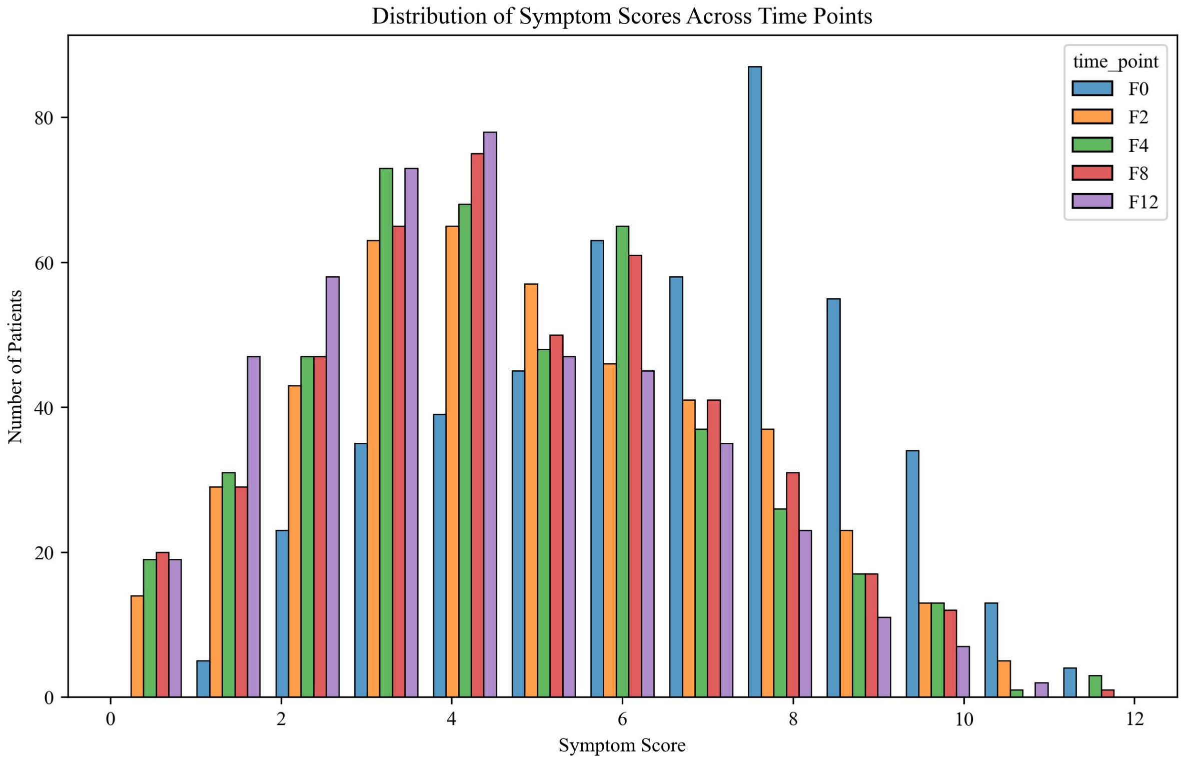


**SFigure 15.** Frequency distribution of symptom scores across study visits. The figure shows the number of participants within each symptom score category at baseline (F0) and follow-up visits (F2, F4, F8, and F12). The distribution gradually shifts toward lower symptom scores over time, indicating reduced symptom burden while maintaining variability between participants.

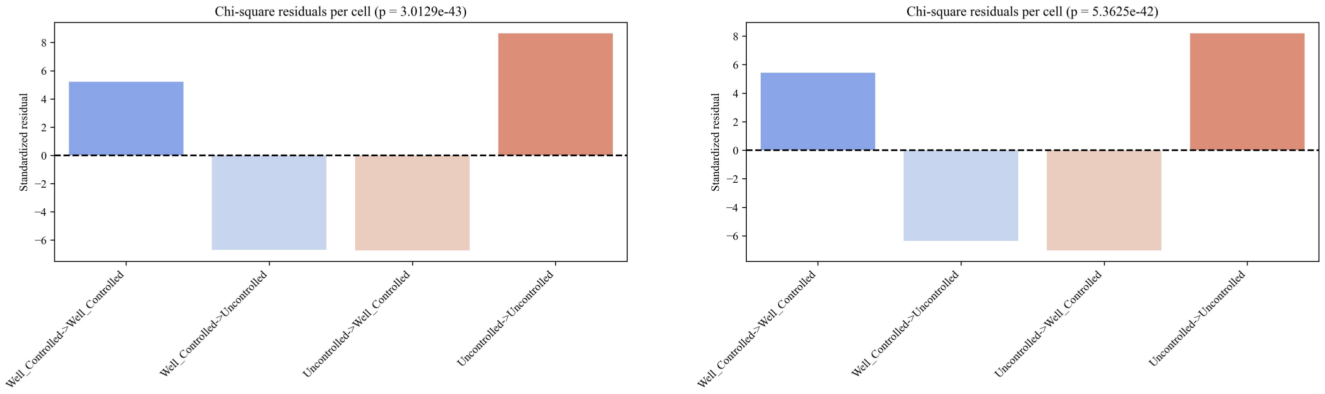


**(a)** Placebo group **(b)** Budesonide group

**SFigure 16.** Standardized Pearson residuals from chi-square tests comparing asthma control status predicted by the miRNA-based Random Forest classifier to observed status (Well-Controlled vs. Uncontrolled). Higher residuals indicate more correct predictions than expected by chance. Associations are highly significant in both groups (budesonide: $p = 1.59 \times 10^{-20}$; placebo: $p = 1.65 \times 10^{-19}$).

### Acknowledgment

This work was supported by the National Institutes of Health (NIH) grants R01 HL162570, R01 HL161362, R01 HL155742, R01 HL177625, and K99 HL183694.
